# Infection Risk Perception, Reporting and Post-Exposure Management of Occupational Injuries Among Healthcare Workers in District Hospitals (Yaoundé, Cameroon)

**DOI:** 10.1101/2023.08.29.23294797

**Authors:** Innocent Takougang, Fabrice Zobel Lekeumo Cheuyem, Jonathan Hangi Ndungo, Emilia Enjema Lyonga, François-Xavier Mbopi-Keou

## Abstract

**Background:** The risk of infection among healthcare workers (HCW) is a looming public health problem worldwide. Developing countries are most affected. The present study aimed to identify knowledge gaps, risk perceptions, reporting patterns, and post-exposure management, following needle stick and sharp injuries and contact with other body fluids HCW.

**Methods:** A cross sectional descriptive study was carried out from January to April 2022 in six District Hospitals in Yaoundé. An auto-administered questionnaire was addressed to consenting HCW. Data were analyzed using R statistic version 4.2.3 and *p*-value<5% was considered significant.

**Results:** Out of the 217 HCWs that were enrolled, some 10% were unaware of sources of exposure to blood and other body fluids (BBF). Respondents displayed insufficient awareness (74-94%) of the main infectious agents that are likely to be transmitted during an accidental exposure to body fluids (AEB). Some HCWs (6,9%) reported a lack of knowledge of the vaccine against Viral Hepatitis B (VHB). Almost half of participants did not know the appropriate recommended action for post-exposure case management (42.9%). Most HCW who had experienced exposure (53.7 %) reported that they did not receive any post-exposure care. Such exposure was often (56.4%) not reported. There was a horizontal variation in underreporting for the same level of healthcare facilities, as it was higher in the Efoulan (OR=3.33) and Nkolndongo (OR=5) District Hospitals. Reasons explaining AEB underreporting were underestimation of vulnerability (51 %) and lack of unawareness of existing post-exposure prophylaxis procedures (42%).

**Conclusion:** While most HCW are aware of the risk of infection associated with exposure to blood and body fluids, reporting was low. Continued educational activities on the risk of infections associated to AEB is warranted. There is a need to implement and scale up structural facilities to enforce measures to prevent exposure, report cases of accidental exposure, ensure post-exposure prophylaxis and counselling.

## Background

The healthcare industry employs about 59.8 million workers worldwide providing health, management and supporting services [1]. The hospital environment is one of the most hazardous work setting and healthcare workers are exposed to biological, chemical, physical and psychosocial hazards [2]. Biological hazards result from accidental exposure to blood and other body fluids (AEB) through needle stick and sharp injuries (NSSI), stings, cuts and splashes that carry infectious agents such as HIV, hepatitis B and C [3]. NSSI and splash reporting initiates the case management process, post-exposure prophylaxis, allows early detection of seroconversion. Adequate post-exposure management decreases HCW’s anxiety. Few healthcare settings (<10%) implement a standard reporting protocol that provides a clear description of post-exposure case reporting and management procedures, where and how HCWs should seek and obtain timely treatment [4]. Concerns about job security, paperwork and loss of time in follow-up are some reasons explaining underreporting of occupation exposure among HCWs. Other reasons for underreporting exposure are low perception of risk, lack of awareness or inadequate enforcement of reporting procedures among HCWs [5]. Lack of the initial training and sensitization among HCW, is a determinant for underreporting needlestick and sharp injuries (NSSI) [6]. In some documented instances, the incidence of NSSIs reporting was 10-fold higher in health facilities bearing standard reporting systems [7,8]. Injury reporting allows for the identification of hazardous devices and initiation of systemic corrective actions. Low levels of reporting are associated with the perception of the resulting injury as mild or severe [9]. Following exposure, HCWs in developing countries are at higher risk of infection, as per the high prevalence of bloodborne pathogens [10]. The prevention of healthcare associated infections relies on pertaining health policies, infection prevention and control practices, immunization, rational use of injections and invasive procedures [5]. The observance of standard precautions has an added value. They include hand washing, systematic use of personal protective equipment (PPE), proper disposal of used needles and sharps [11]. Settings with low adherence to standard precautions, insufficient training, economic barriers and poor supplies of PPE were associated with high prevalence of occupational exposure [3,12,13]. Despite the body of knowledge on the magnitude of the healthcare associated infections among HCWs, corrective policy measures are often patchy or neglected [8,9]. There is a need to document HCW and health facilities factors that facilitate post-exposure reporting and case management. Such data will be useful in designing intervention strategies to guide the implementation of safety standards in district health facilities.

## Methods

We conducted an institution-based cross-sectional study from January to April 2022, covering all District Hospitals of Yaoundé, the political capital of Cameroon.

The Cameroonian health system is organised around health districts. The District Hospital is the first level of reference in the health system pyramid. It is responsible for providing primary health care to a standard population of 10 000 people [14]. The study covered all the six District Hospitals of Yaoundé including Biyem-Assi, Cite-Verte, Djoungolo, Efoulan, Mvog-Ada and Nkolndongo District Hospitals. These health facilities cover a population of 3.2 million residents with 400 health workers and 330 beds. They offer 153 583 consultations and 19 092 hospital admission per annum [13,15].

Study participants were workers who come into contact with patients in their daily tasks, thus potentially exposed to body fluids. They were medical (physician and intern), paramedical (nurse, assistant nurse, midwife, laboratory and dental technician) and hygiene (cleaners, sanitation engineer) workers.

The sample size was calculated using the single proportion formula (n = [z_α/2_]^2^ *[p (1-p)]/d^2^), where z_α/2_ = 1.96 and p=37 % was obtained from a similar study in referral hospital in Yaoundé [16]. The standard error was d=5%. Using above formula, we obtained a minimum required sample size of 198 was obtained. In each clinical department, all consenting personnel were included.

The study tool was a self-administered questionnaire that addressed issues related to sociodemographic and professional characteristics, experience of accidental exposure, reporting and management practices following accidental exposures to blood or body fluids

Data were cross-checked, recoded, entered and analysed using R statistics Version 4.2.3. The Chi^2^ and Fisher’s exact tests were used to compare proportions. Simple binary logistic regressions were used to assess the strength of the association between variables. *p*-value < 0.05 was considered statistically significant.

## Results

### Sociodemographic characteristics

Out of the 279 HCWs that were contacted, 217 returned the completed questionnaire, for a response rate of 77.8 %. Study participants were medical, paramedical and hygiene workers[3,13].

### Knowledge of health risks associated to AEB

Nearly 10 % of participants were unaware of the AEB as a likely source of infection. The lowest levels of awareness were found among paramedics (7 %) and hygiene personnel (40 %) (*p*-value < 0.0001). Half of the hygiene and sanitation staff (50 %) did not know the activities that could lead to exposure (*p*-value < 0.0001) (Table I).

**Table I:**
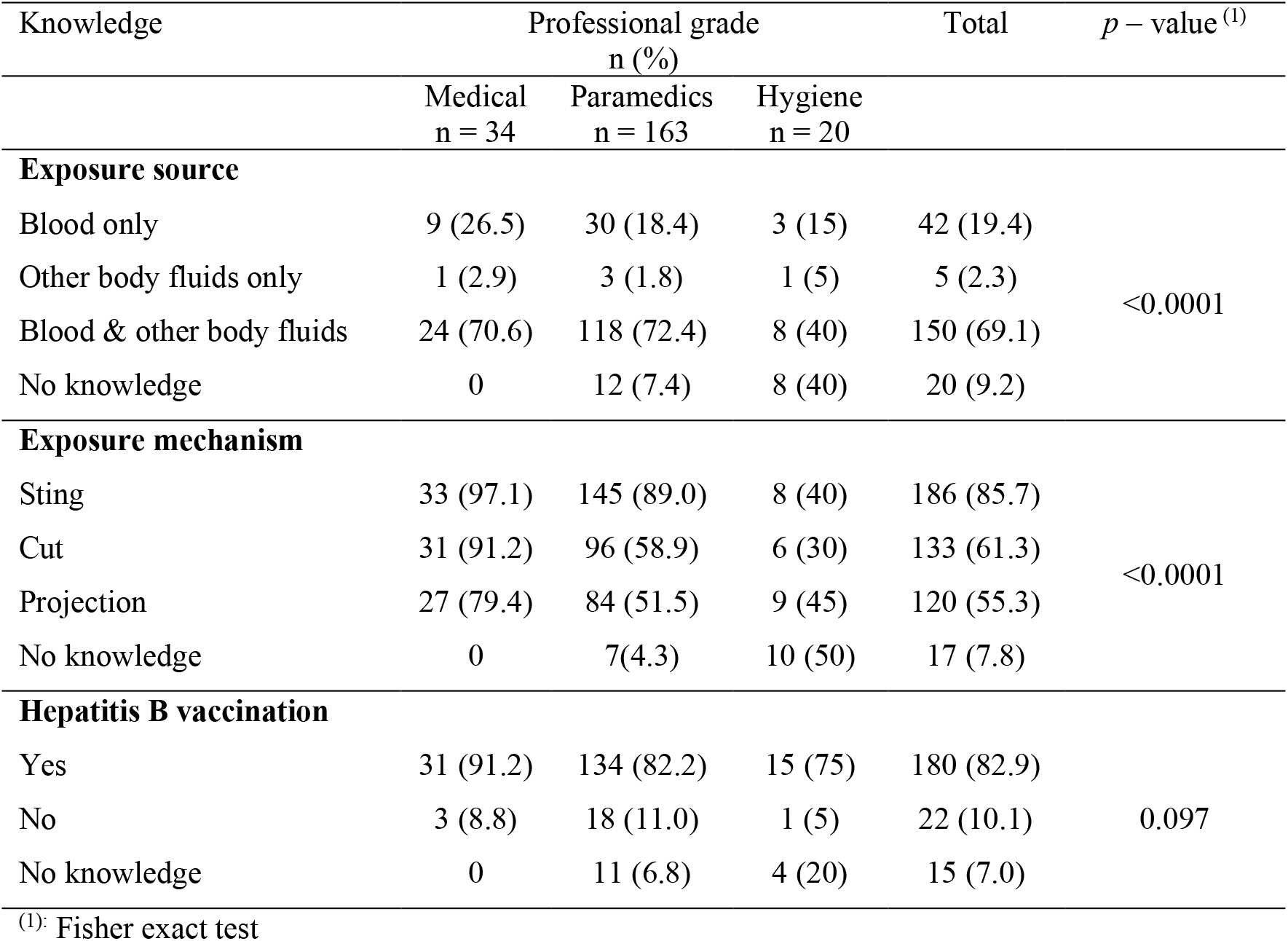
Awareness of health risk of Occupational Exposure to Blood and other Body fluids among Healthcare Workers in Yaoundé District Hospitals, April 2022 (*n=*217)

Most HCW were aware that the blood-borne pathogens HIV, hepatis B and C could be transmitted through AEB. Most respondents were aware that the SARS-COV-2 could be transmitted in the course of an AEB (99.5%) (Figure 1).

**Figure 1:**
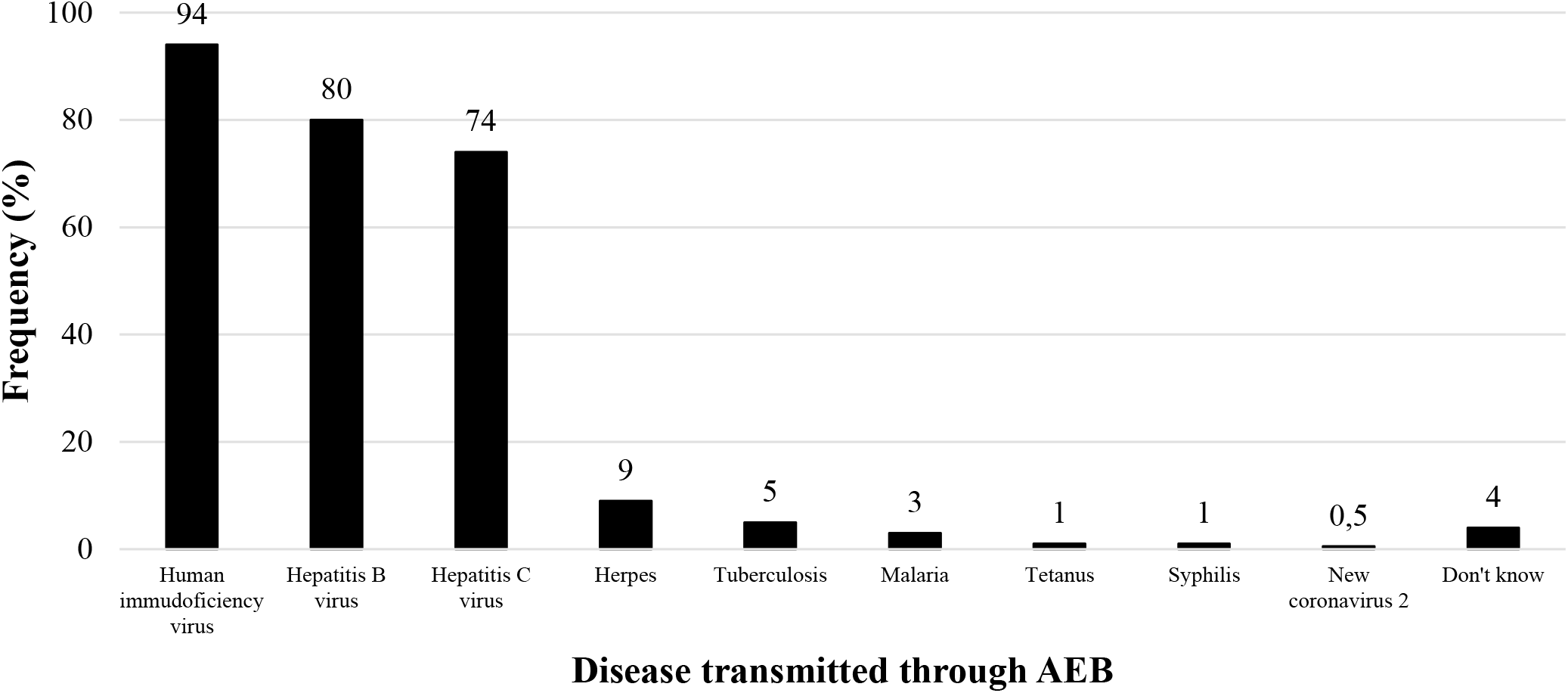
Healthcare Workers Awareness of Infectious agents likely to be Transmitted through Accidental Exposure to Blood and other Body fluids, Yaoundé District Hospitals,

More than half of HCW were aware of the first recommended action in case percutaneous exposure to blood (57.1 %) (Table II)

**Tableau II:** Knowledge of Emergency Action in the Post-Exposure Management among Healthcare Workers in Yaoundé District Hospitals.

| Emergency action | Professional grade<br>n (%) |  |  | Total | p – value |
| --- | --- | --- | --- | --- | --- |
|  | Medical<br>n = 34 | Paramedics<br>n = 163 | Hygiene<br>n = 20 |  |  |
| Let the wound bleed | 9 (26.5) | 54 (33.1) | 2 (10.0) | 65 (30.0) | 0.083<sup>(2)</sup> |
| **Wash thoroughly with soapy water** | 20 (58.8) | 98 (60.1) | 6 (30.0) | 124 (57.1) | 0.037<sup>(1)</sup> |
| Apply alcohol only | 6 (17.6) | 5 (3.1) | 5 (25) | 12 (5.5) | 0.006<sup>(2)</sup> |
| Apply bleach only | 2 (5.9) | 11 (6.7) | 5 (25.0) | 18 (8.3) | 0.037<sup>(2)</sup> |
| Don't know | 1 (3.0) | 0 | 1 (5) | 2 (0.9) | 0.557<sup>(2)</sup> |

<sup>(1)</sup>: Chi<sup>2</sup> test; <sup>(2)</sup>: Fisher exact test

### Reported emergency actions in post-exposure care

After a percutaneous exposure to blood, HCW washed the wound with soapy water (86%) or with running water (44 %). Some squeezed and bled the affected site (32%), while few respondents did not do anything (3%) (Figure 2).

**Figure 2:**
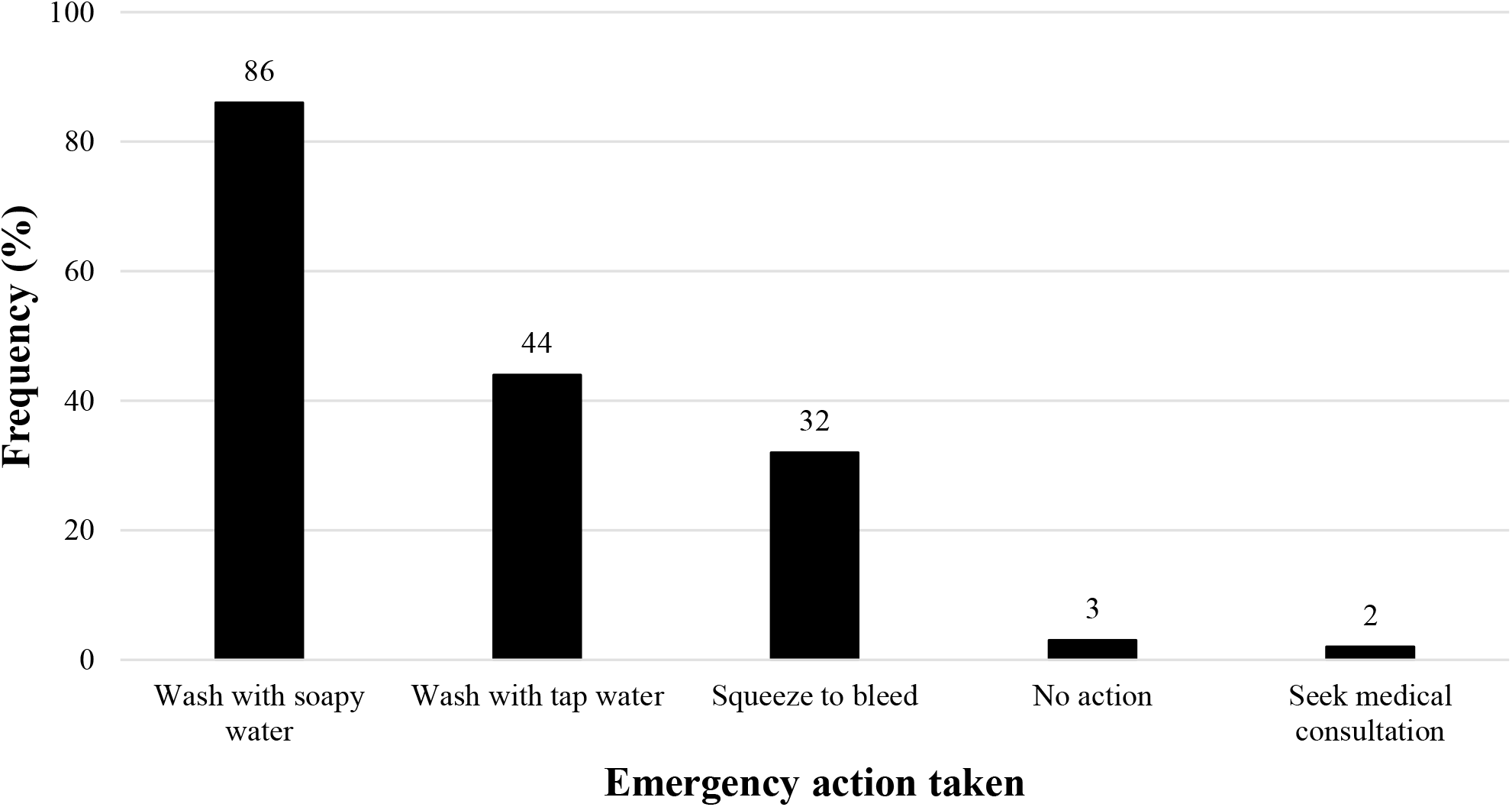
Reported first Action after Exposure to Blood among Healthcare Workers in Yaoundé District Hospitals,

More than half of the affected healthcare workers (53.7 %) reported that they did not receive any post-exposure care. Post exposure prophylaxis (69 %) and HIV screening (45 %) were the most reported post-exposure care received by affected healthcare workers (Figure 3).

**Figure 3:**
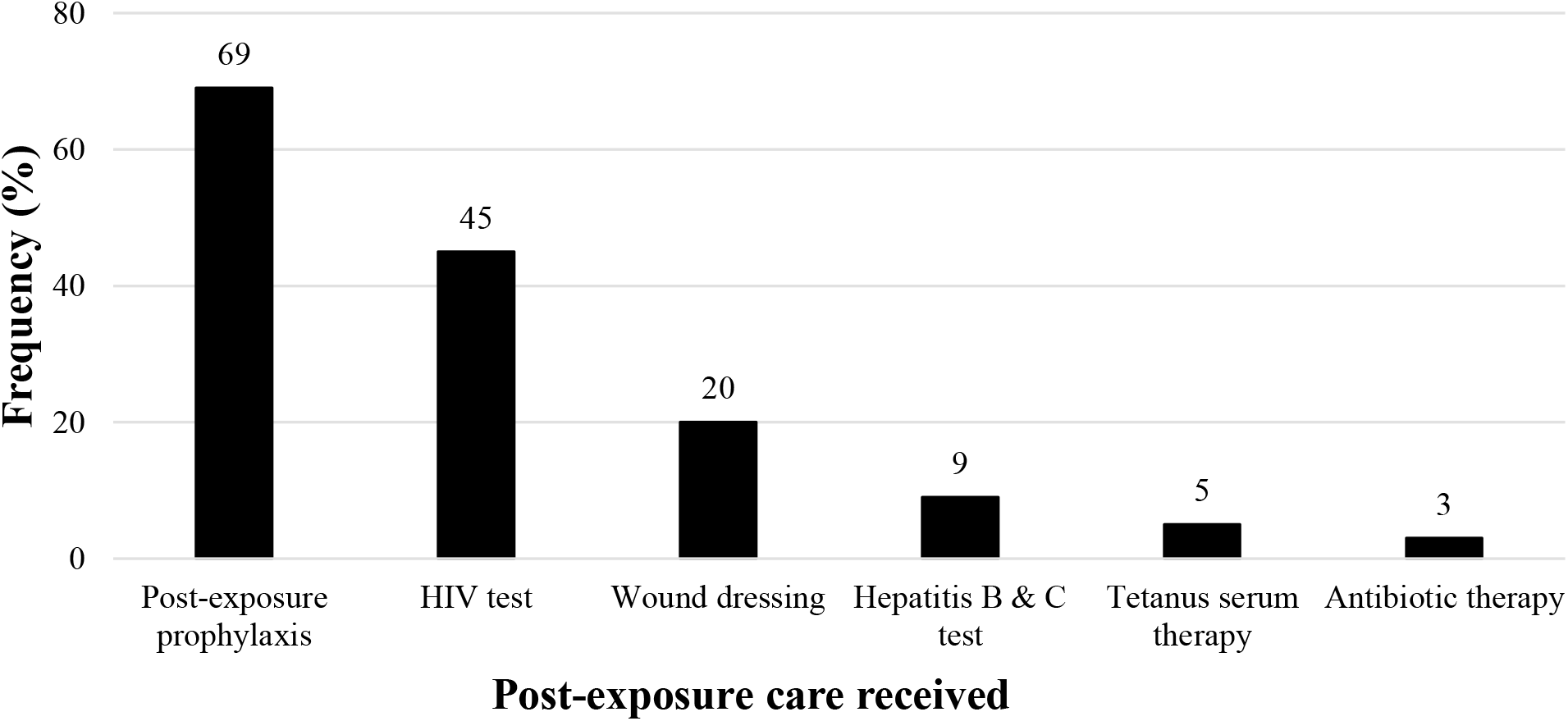
Post-exposure Care received following Work-related Injuries among Healthcare Workers in Yaoundé District Hospitals, April 2022 (*n* = 69)

### Occupational exposure reporting

Almost all healthcare workers (94.9 %) were aware that AEB should be reported. Out of the 149 participants that had experienced percutaneous injury, more than half (56.4%) did not report the last exposure they suffered. More than three quarters (77%) did not know the personnel in charge of the post-exposure case management in the study health facilities.

Taking the Cite-Verte District Hospital as a reference, the risk of underreporting was significantly higher in the Efoulan (OR=3.33) and Nkoldongo (OR=5) (Table III).

**Table III:**
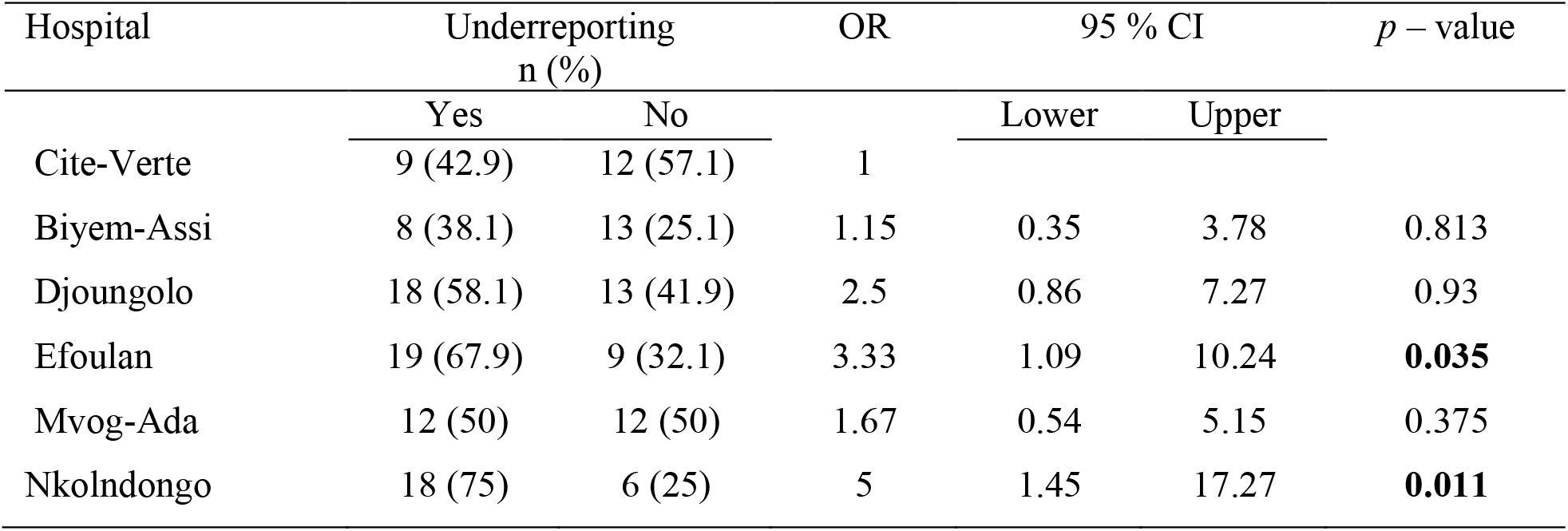
Simple Binary Logistic Regression of the Risk of Underreporting among Healthcare Workers in Yaoundé District Hospitals.

| Hospital | Underreporting<br>n (%) |  | OR | 95 % CI |  | *p* – value |
| --- | --- | --- | --- | --- | --- | --- |
|  | Yes | No |  | Lower | Upper |  |
| Cite-Verte | 9 (42.9) | 12 (57.1) | 1 |  |  |  |
| Biyem-Assi | 8 (38.1) | 13 (25.1) | 1.15 | 0.35 | 3.78 | 0.813 |
| Djoungolo | 18 (58.1) | 13 (41.9) | 2.5 | 0.86 | 7.27 | 0.93 |
| Efoulan | 19 (67.9) | 9 (32.1) | 3.33 | 1.09 | 10.24 | **0.035** |
| Mvog-Ada | 12 (50) | 12 (50) | 1.67 | 0.54 | 5.15 | 0.375 |
| Nkolndongo | 18 (75) | 6 (25) | 5 | 1.45 | 17.27 | **0.011** |

Out of the 84 HCW who did not report the last percutaneous exposure, half (51 %) explained that the risk of contamination was low, or they were unaware of post-exposure procedures (42 %) (Figure 4).

**Figure 4:**
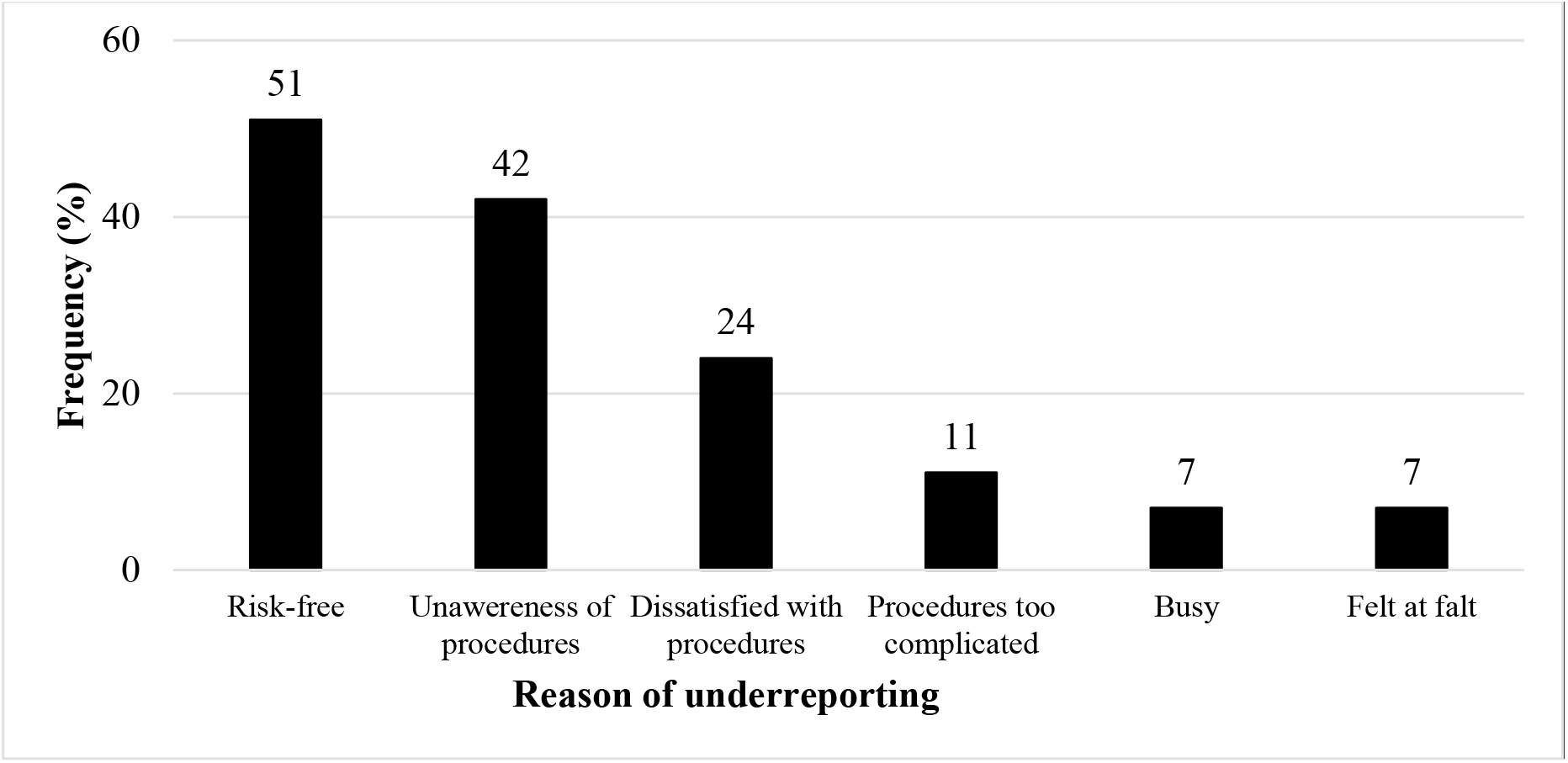
Reported Reasons of Underreporting Exposure to Body Fluids among Affected Healthcare Workers in Yaoundé District Hospitals, April 2022 (*n* = 84)

## Discussion

### Risk perception

Health staff knowledge assessment on AEB found that almost a third of respondents could not identify all sources of contamination during AEB. This knowledge gap was statistically significant among hygiene staff. This would be due to the low educational level within this particular professional group and the lack of training on the topic.

HCW had good knowledge on AEB mechanisms of occurrence during healthcare activities. Nevertheless, nearly 10 % had no knowledge of these mechanisms and half of them were hygiene staff. Similar findings were observed in Congo and Togo [2,17]. Stings from used sharp objects were the mechanism frequently identified by HCW in most studies [2,18,19].

Most of HCW were aware of blood-borne pathogens (HIV, hepatis B and C) transmitted through AEB. The priority identification of HIV would be due to the fact that this blood-transmissible virus and its derivatives is well known because of its global health consequences in general and particularly in Sub-Saharan Africa [18]. Our result corroborates observations in other health settings in Cameroon, Congo and Morocco [2,16,18].

Most of HCW were aware of protection provided by vaccination against viral hepatitis B. The prevalence of viral hepatitis B is considered high (11.2 %) in Cameroon and could arouse among HCW more interest on vaccination which is one the cost effective means of prevention [20].

Washing the wound with water and soap is the immediate action recommended after percutaneous exposure to BBF. Almost half of the HCW were not aware of this recommendation. Similar result was obtained in Congo [21]. The lack of continuous training on AEB could explain this poor adherence to related guidelines. In France, nurses who had received education on AEB were able to report it accurately, while those who had only practical experience expressed a strong need for information and awareness. The most at ease were those who had benefited from a well-supervised internship and lectures integrated into their academic curriculum [18].

### AEB Reporting and Management

Most of HCW were aware of the need to report AEB to whom it may concern corroborating findings in China [4]. However, our study found that more than half did not report the last percutaneous exposure suffered. Similar results were obtained in Ethiopia [5,22]. Outlined reasons explaining this high proportion of underreporting included the perception that the patient care for was risk free, unawareness of post-exposure procedures and dissatisfaction with procedures among others. Our results corroborate observations in Kenya and Ethiopia [5,22,23].

The horizontal variation in underreporting patterns among health facilities of the same hierarchical level suggests that specific need for education and information of staff depends on the health facilities. Moreover, the absence of information and the underestimation of the risk related to percutaneous exposure therefore appeared to be the reasons explaining the underreporting. Nkoldongo, Djoungolo and Efoulan District Hospitals should take more measures so that their employees refer to the person in charge of the management of the AEB who is most often the nurse officer of HIV service. To this end, provisions should be made so that they are known to all staff in order to encourage and facilitate the reporting process because more than three quarters of HCW did not know the manager in charge of the AEB in those health facilities. Similar findings were obtained in Nigeria and Morocco [18,24]. From a medico-legal point of view, any suspicious injury occurring as a result of or in the course of work is covered as an accident at work or occupational disease and requires a compensation as it is the case in Morocco [18]. A similar health policy should be put in place in Cameroon to decrease the burden of AEB and related consequences on HCW health.

Most of HCW stated they washed the affected site with soapy water [25,26]. Our results corroborates findings in two reference hospital-based studies in Cameroon and Kenya [16,23]. On the other hand, A third of participants had let the wound bleed after exposure. Studies conducted elsewhere in Kenya and Ethiopia found lower proportions [5,23]. According to the recommendations available in some studied District Hospitals [13] and enacted by Cameroon health authorities, letting the wound bleed after a percutaneous exposure have no influence on the transmission of blood-borne pathogens. This recommendation, however, remains controversial because some authors believe that letting the wound bleed is an immediate action in case of percutaneous exposure [27].

Most of HCW reported, they received a short course of antiretroviral therapy (69%) after percutaneous exposure. This suggests the contaminating nature of the exposures suffered. However, this proportion was higher than those obtained in referral hospital settings in Cameroon, Kenya and Ethiopia [5,16,23]

Less than half of HCW had benefited from HIV screening. Poor compliance with procedures, likely due to fear of loss of confidentiality of serological status, or shortcuts of screening inputs could explain this low screening rate. Our result corroborates finding in other health facilities in Cameroon and Ethiopia [5,16].

## Conclusion

This study revealed a good perception of disease and need to report accidental exposure to blood among healthcare workers. However, poor administrative standards seemed to have influenced the reporting and proper management of AEB in Yaounde health facilities. Therefore, more rigorous control measures for occupational injuries prevention and their management need to be implemented by national health authorities.

## Data Availability

All data produced in the present study are available upon reasonable request to the authors

https://doi.org/10.29011/2577-2228.100321

http://dx.doi.org/10.34297/AJBSR.2023.19.002628

## Declaration

### Author’s Contribution

Drafting of the study protocol, data collection, analysis and interpretation, drafting and editing of manuscript: F.Z.L.C.; Critical revision of protocol, critical revision of manuscript: J.H.N., E.E.L., and M-K. F-X.; Conception, design and supervision of research protocol and implementation, data analysis plan, revision, editing and final validation of the manuscript: I.T.

### Ethical Approval Statement

The protocol was reviewed and approved by the Regional Human Health Committee of the Centre (CRERSH-Ce) and the ethical clearance: CE N° 2245/CRERSHC/2021 issued.

## Acknowledgements

Our gratitude goes to the health personnel who agreed to participate in this study and to the managers of the health facilities who gave their authorisation for the conduct of this study.

